# Monitoring Italian COVID-19 spread by an adaptive SEIRD model

**DOI:** 10.1101/2020.04.03.20049734

**Authors:** Elena Loli Piccolomini, Fabiana Zama

**Affiliations:** Department of Computer Science and Engineering, University of Bologna, Italy; Department of Mathematics, University of Bologna, Italy

## Abstract

Due to the recent diffusion of COVID-19 outbreak, the scientific community is making efforts in analysing models for understanding the present situation and predicting future scenarios. In this paper, we propose a Susceptible-Infected-Exposed-Recovered-Dead (SEIRD) differential model [Weitz J. S. and Dushoff J., Scientific reports, 2015] for the analysis and forecast of the COVID-19 spread in Italian regions, using the data from the Italian Protezione Civile from February 24th 2020. In this study, we investigate an adaptation of SEIRD that takes into account the actual policies of the Italian government, consisting of modelling the infection rate as a time-dependent function (SEIRD(rm)). Preliminary results on Lombardia and Emilia-Romagna regions confirm that SEIRD(rm) fits the data more accurately than the original SEIRD model with constant rate infection parameter. Moreover, the increased flexibility in the choice of the infection rate function makes it possible to better control the predictions due to the lockdown policy.

## Introduction

The recent diffusion of the COVID-19 Corona-virus has renewed the interest of the scientific and political community in the mathematical models for the epidemic. Many researchers are making efforts for proposing new refined models to analyse the present situation and predict possible future scenarios.

With this paper, we hope to contribute to the ongoing research on this topic and to give a practical instrument for a deeper comprehension of the virus spreading features and behaviour.

The modelling of infectious diseases is currently performed by Ordinary Differential Equations (ODEs) deterministic compartmental models [1] or by stochastic procedures [2]. The tuning of the parameters of the equations allows better modelling of environmental features, such as social restrictions or changes of political strategies in the outbreak containment.

We consider here deterministic compartmental models, based on a system of initial value problems of Ordinary Differential Equations. This theory has been studied since the beginning of the century by W.O. Kermack and A. G. MacKendrick [3] who proposed the basic Susceptible-Infected-Removed (SIR) model. The SIR model and its later modifications, such as Susceptible-Exposed-Infected-Removed (SEIR) [4] were later introduced in the study of outbreaks diffusion. These models split the population into groups, compartments, and reproduce their behaviour by formalising their reciprocal interactions. For example, the SIR model groups are Susceptible who can catch the disease, Infected who have the disease and can spread it, and Removed those who have either had the disease or are recovered, immune or isolated until recovery. The SEIR model proposed by Chowell et al. [5] also considers the Exposed group: containing individuals who are in the incubation period.

The evolution of the Infected group depends on a critical parameter, usually denoted as R0, representing the basic reproductive rate and it measures of how transferable a disease is. This quantity determines whether the infection will spread exponentially, die out, or remain constant. When *R*_0_ *>* 1 the epidemic is spreading. The value of R0 can be inferred, for example, by epidemic studies or by statistical data from literature or it can be calibrated from the available data. In this paper, we use the available data for determining the value of R0.

Compared to previous outbreaks, such as SARS-CoV or MERS-CoV [6], when the disease stopped after a relatively small number of infected people, we are now experimenting a completely new situation, and the COVID-19 epidemic has been proclaimed pandemic by the WHO. Indeed, the number of infected people grows exponentially, and apparently, it can be stopped only by a complete lockdown of the affected areas, as evidenced by the COVID-19 outbreak in the Chinese city of Wuhan in December 2019.

Analogously, in the Italian case, to limit the virus diffusion all over the Italian area, the government has started to impose more and more severe restrictions since March 10th 2020. Hopefully, these measures affect the spread of the COVID-19 virus, reducing the number of infected people and the value of the parameter R0.

The introduction of different levels of lockdown requires an adaptation of the standard epidemic models to this new situation, see [6–8] for some example related to the Chinese outbreak. Concerning the Italian situation, the outbreak started in Lombardia on February 21st, and it is still affecting more and more regions. In the beginning, severe lockdown measures were imposed in very restricted areas and only after March 10th uniform restrictions were imposed all over the country.

We believe that a more flexible, time-dependent parameter related to the infection rate, not only could it give a better fit of the measured data, but it also allows us to represent possible future scenarios caused by different policies of the Italian Government. In this paper, we propose to represent the infection rate as a piecewise time-dependent function, which reflects the changes in external conditions. The parameter R0, which is proportional to the infection rate, becomes a time-dependent parameter *R*_*t*_ and it follows a different trend each time the external conditions change, depending on the particular situation occurring in that period. For example, the application of new restrictions to the population movements at time *t*_0_ should hopefully cause a decrease of *R*_*t*_ when *t > t*_0_.

The idea of modelling the introduction of restricting measures through a non-constant infection rate has been proposed in [5], where Ebola data coming from Congo and Nigeria are analysed. Although the particular form of time-dependent infection rate, used in [5], gave outstanding results in that particular situation, it is not efficient in our case. Therefore, we propose a different function type for COVID-19 diffusion in Italy. We split the observation domain into several sub-intervals and represent the infection rate as a piecewise rational function.

Moreover, we believe that relevant information is not only about Infected but also Recovered and Dead numbers, hence we choose to apply the SEIRD, model by splitting the Removed population into Recovered and Dead.

Finally, some considerations on how to use the proposed model and about the effectiveness of the present research. The COVID-19 data, currently available in Italy, is quite raw and incomplete, due to the present emergency. Appropriate use and interpretation require data preprocessing. For example, apparently, all the dead people positive to COVID-19 are counted as *Dead*, but probably many of them might have died in a few days for other diseases. For this reason, the model results, although coherent with the available data, may not depict the Italian situation accurately. Of course, the proposed model could be easily applied to new and possibly improved data.

Concerning the prevision obtained by applying the proposed model for times beyond the measured interval, we remind that, as for all forecasts, the more considerable the distance is from the observed data, the more unreliable is the forecast. However, the model flexibility allows us to adapt it to changes in the data trends.

The paper is organized as follows. In section 1, we describe the details of SEIRD model with constant and with time-dependent infection rate SEIRD(rm). Finally in section 2 we test the model on some regional aggregated data published by the Protezione Civile Italiana [9].

## 1 Materials and methods

In this section we present the proposed mathematical model for the COVID-19 analysis in Italy, the method used for estimating the model parameters by data fitting and the strategies applied for predictions.

### The proposed SEIRD model

The epidemiological compartmental model divides the population into groups, whose evolution in time is described by continuous functions, and model the relations between the groups with ODEs involving the relative functions. The first differential model proposed in literature is the SIR model by Kermack and A. G. MacKendrick [3], considering the groups of Susceptible (S), constituted by people that can be infected, Infectious (I) and Removed(R), containing recovered and dead people. The more recent SEIR model adds the Exposed (E) group to the previous one, representing people that infected but not infectious. SEIR has been used to model breakouts, such as Ebola in Congo and Uganda [4, 5]. In [4] the equations are modified by adding the quarantine and vaccination coefficients. In our case, unfortunately, vaccination is not available.

In this paper we use a SEIRD model [10], where the class of Removed in the SEIR model is partitioned into the Recovered (again labelled with (R)) and Dead (D). Hence SEIRD consider five classes: Susceptible (S), Exposed (E), Infectious (I), Recovered (R) and Dead (D). Their sum, at each time t, is the total number N of individuals in the examined population, i.e. *N* = *S* + *E* + *I* + *R* + *D*. The system of equations in the SEIRD model is given by:

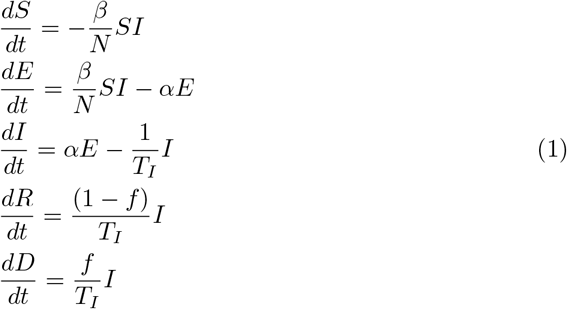

where *N* is the total population, *β* is the infection rate, a coefficient accounting for the susceptible people get infected by infectious people, *α* represents the number of days for the transition from Exposed to Infectious (i.e. the incubation rate), *T*_*I*_ is the average infectious period and *f* is the fraction of individuals who die.

The system (1) of ODEs is solved by starting from an initial time *t* = *t*_0_ where the values of the populations *S*(*t*_0_), *E*(*t*_0_), *I*(*t*_0_), *R*(*t*_0_), *D*(*t*_0_) are assigned on the basis of the available data at that time and integrated up to a final time *T*.

Following [10], we compute the basic reproduction number *R*0 as follows:

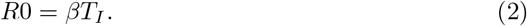

In order to consider different external conditions affecting the outbreak data, we partitioned the integration interval [*t*_0_, *T*] into sub-intervals [*t*_*k*_, *t*_*k*+1_], *k* = 1, … *p* and, in each sub-interval, we adapt the model to the data. Possible conditions changing the outbreak diffusion trend are the different restrictions imposed by the Italian government since March 10th 2020, or different protocols regarding swabs. Since the applied restrictions should cause a decrease in the number of contacts between Infected and Susceptible, we model the coefficient *β* in (1) as a decreasing time-dependent function *β*(*t*).

A similar model for the infection rate in SEIR equations can be found in [5], where the function is assumed to have a decreasing exponential form. However, observing the data trend, we have proved that the exponential *β*(*t*) has a too fast decreasing behaviour and we choose to model it as a decreasing rational function:

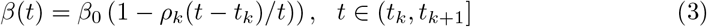

where *ρ*_*k*_ ∈ (0, 1) for *k >* 0 and *ρ*_0_ = 0. The introduction of *β*(*t*) (3) in the SEIRD model (1) originates the SEIRD *r*ational *m*odel (SEIRD(rm)) reported below,

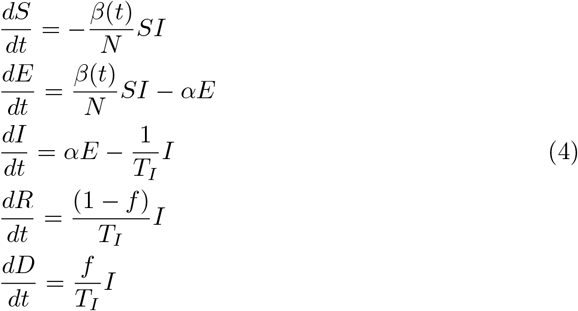

since infection rate *β*(*t*) is time-dependent, the the basic reproduction number is written as follows:

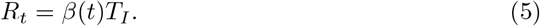

### Parameter estimation

We calibrate the parameters of SEIRD (1) and SEIRD(rm) (4) equations by solving non-linear least squares problems with positive constraints. For example, in the SEIRD model (1), we define the function **u**(*t*) = (*S*(*t*), *E*(*t*), *I*(*t*), *R*(*t*), *D*(*t*)), depending on the vector of parameters **q** = (*β, α, f*), and the vector **y** of the acquired data at given times *t*_*k*_, *k* = 1, … *p*. Let *F* (**u, q**) be the function computing the numerical solution **u** of the differential system (1), the estimation of the parameter **q** is obtained solving the following non linear least-squares problem:

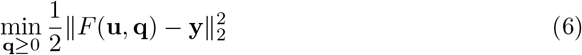

where we introduce positivity constraints on **q**. The trust-region based method implemented in the lsqnonlin Matlab function is applied to solve problem (6) (see [11] for details about this aspect).

Actually, we implement the parameter identification step as a two phases process:

- **Phase 1** Identification of the parameters (*α, β, f*) in (1), using a restricted data set: in our examples we considered the data of the first 18 days.
- **Phase 2** Identification of the parameters (*α, f*) in SEIRD(rm)(4) model, using all the measures available (times *t*_0_, *t*_1_, …, *t*_*p*_) and modelling the infection rate as in (3). This second phase uses the values of the parameters of Phase 1 as starting guess, and sets *β*_0_ in (3) as the value *β* estimated in Phase 1.

### Prediction

After having carried out the estimation of the parameters of (4), based on the data available until March 26 2020, we use the model to obtain some predictions about the infection behaviour in the successive few weeks. This information is extremely important to evaluate if the restrictions give the expected results and to predict the length of the epidemic spread.

Concerning the forecasts, we propose three possible ways to model the infection rate for the subsequent times where the data is still not available. We extend the computation domain by adding the interval [*t*_*p*_, *t*_*s*_], where *t*_*p*_ is the time value of the last measure and *t*_*s*_ is the final simulation time. Then we solve equation (4) by applying (3) for *t* ∈ [0, *t*_*p*_] and representing *β*(*t*) in one of the following three different ways:

1. piecewise constant function *β*_*c*_(*t*):

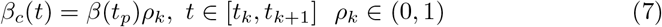
2. piecewise rational function *β*_*r*_(*t*):

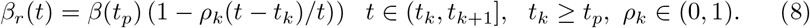
3. piecewise exponential function *β*_*e*_(*t*):

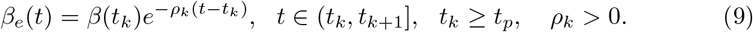

By changing the number of partition points *t*_*k*_ of the simulation domain and the weights *ρ*_*k*_, for each formula it is possible to take into account the different restriction measures and estimate possible developments of the epidemic, such as the behaviour of the Infected, Recovered or Dead individuals.

## 2 Results

In this section, we apply the SEIRD (1) and the SEIRD(rm) (4) models to monitor the Covid-19 outbreak in Italy during the period 24/02/2020-27/03/2020. The epidemic spread started on February 21st affecting the northern regions. Lombardia in particular registered the first sources of epidemic followed by the Veneto and Emilia-Romagna.

Since initially each region applied different containment measures to some restricted areas, at different times, we choose to calibrate the SEIRD and SEIRD(rm) models in each region separately. This study considers two regions, Lombardia and Emilia-Romagna, for which the available data can be found in the GitHub repository [9].

All the computations are performed using Matlab R2019b 2,9 GHz Intel Core i7 quad-core 16 GB ram. We separately present the results of the two steps described in section 1:

**Identification**. This step computes estimates of the model parameters using a set of measured data and solving the minimization problem (6).
**Simulation**. This step applies the SEIRD(rm) (4) model, with the identified parameters, to monitor the epidemic evolution in subsequent times. Since currently the epidemic has not still reached the peak of infection, we test different types of variable infections rates (7), (8), (9) to predict the possible evolution.

The differential systems SEIRD and SEIRD(rm) are solved applying the ode45 Matlab function, implementing a variable step Runge-Kutta method based on Dormand-Prince formulae, with the following initial condition: *S*(*t*_0_) = *N, E*(*t*_0_) = *I*(*t*_0_) = *I*_*init*_, *R*(*t*_0_) = *D*(*t*_0_) = 0, where the value *I*_*init*_ corresponds to the Infected individuals in the first measurement day and *N* is the total population of the region. From the database in [9], we considered the Infected individuals in the 11th column (the given number of total positive cases).

The average infectious period, obtained from data estimations as well as from clinical observation, is assumed as *T*_*I*_ = 20.

To test the precision of our data estimation process, we consider the following relative error between the vector *X*_*mod*_ of the modelled data and the vector *X*_*data*_ of the the measured data for the days 1, 2 … 31:

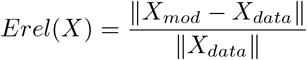

where *X* represents the considered model compartment {S,E,I,R,D} respectively, and ∥· ∥ is the Euclidean distance.

### 2.1 SEIRD calibration

In this paragraph we test the SEIRD model (1) to estimate the model parameters **q** = (*β, α, f*) twice, first by using the data in [9] from 18 days (24/02/2020-12/03/2020) and then by using the data of all the 31 days (24/02/2020-27/03/2020), respectively.

From the values reported in table 1 we observe that the parameters obtained from 31 days mostly lead to worse errors, compared to those obtained from the first 18 days. Moreover, observing the plots in figures Fig. 1, Fig.2, Fig.3 and Fig.4, we conclude that SEIRD model does not seem to reproduce accurately the slopes of the Infected, Recovered and Dead data; hence any prevision based on this model is not reliable. The reason is that, since the data trend is not the same for the whole period of 31 days, a constant value for the *β* parameter, i.e. for the transmission rate, is not correct. However, the computed parameters reported in table 2 can be used a starting guess for the SEIRD(rm) model at Phase 2.

**Fig 1.**
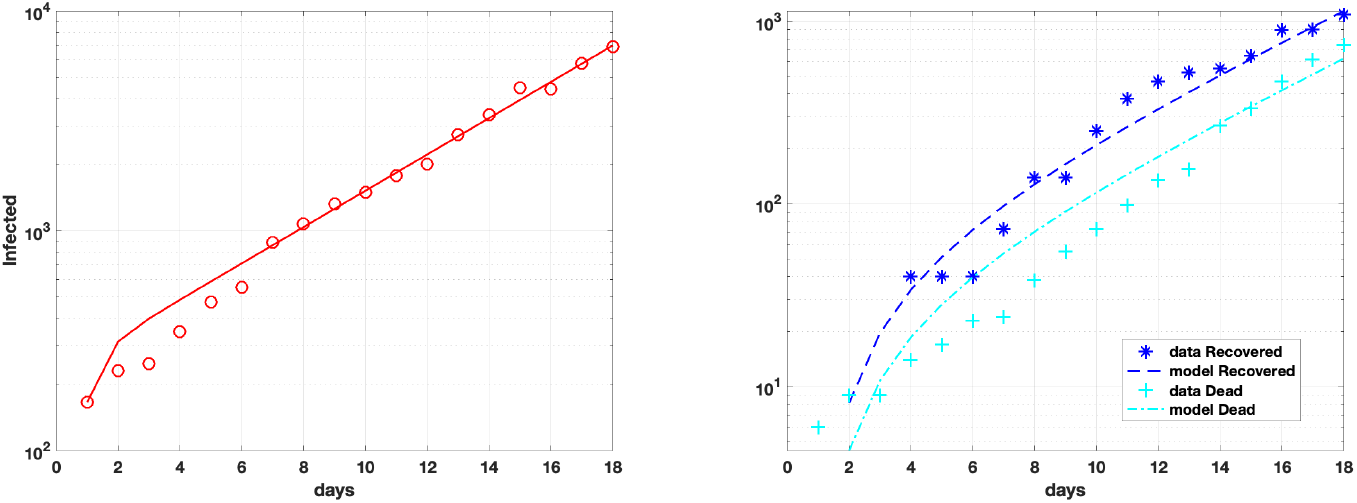
Infect, Recovered and Dead fit based on Lombardia data 18 days.

**Fig 2.**
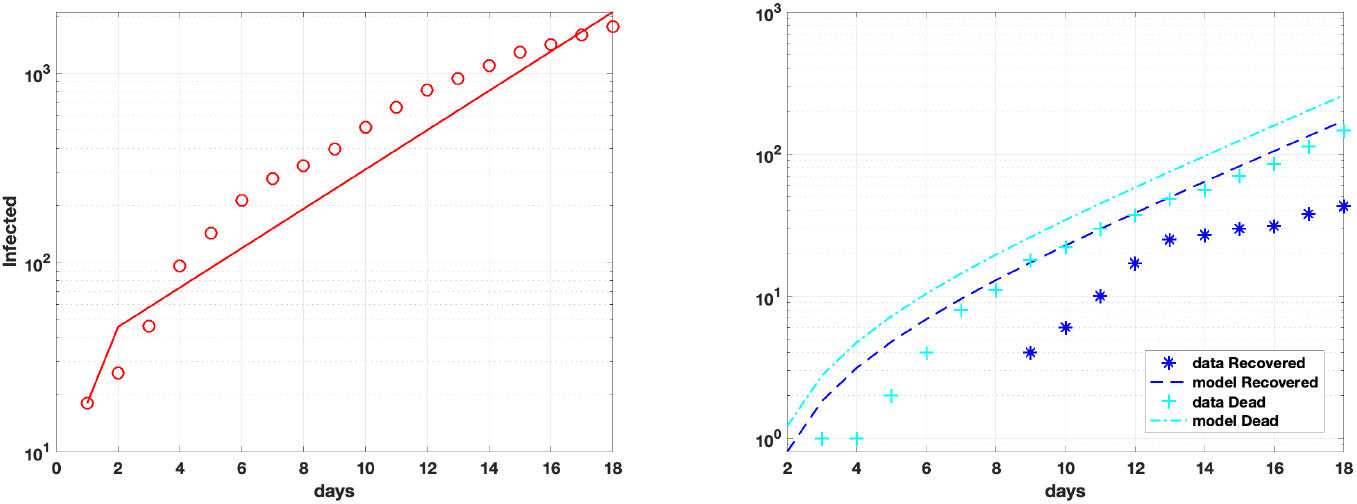
Infect, Recovered and Dead fit based on Lombardia data 31 days.

**Fig 3.**
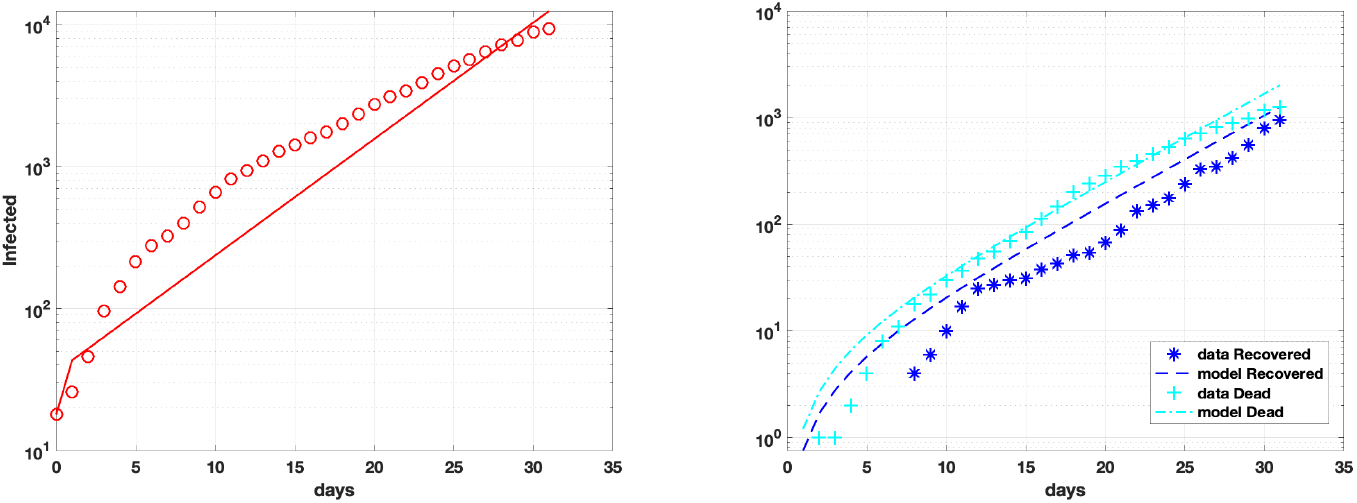
Infect, Recovered Dead fit based on Emilia Romagna data 18 days.

**Fig 4.**
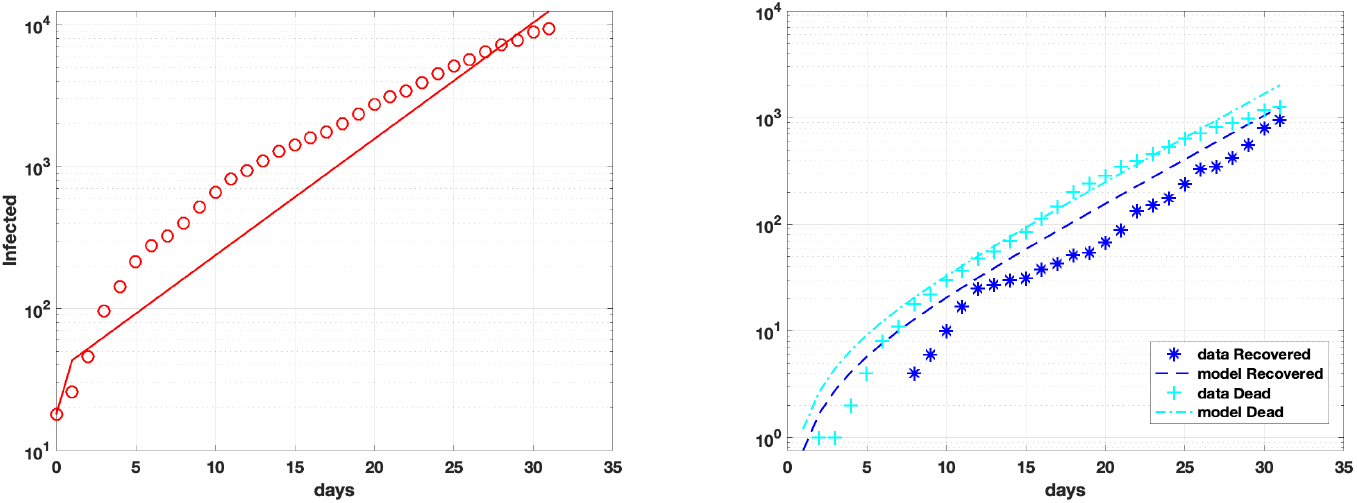
Infect, Recovered Dead model fit on Emilia Romagna data 31 days.

**Table 1.** SEIRD Relative Errors in two different time intervals: 18 days (24/02/2020-12/03/2020); 31 days (24/02/2020-27/03/2020).

| Region | N days | $Erel(I)$ | $Erel(R)$ | $Erel(D)$ |
| --- | --- | --- | --- | --- |
| Lombardia | 18 | $6.17 \cdot 10^{-2}$ | $1.33 \cdot 10^{-1}$ | $1.76 \cdot 10^{-1}$ |
| | 31 | $2.33 \cdot 10^{-1}$ | $2.67 \cdot 10^{-1}$ | $4.32 \cdot 10^{-1}$ |
| . Emilia R. | 18 | $2.24 \cdot 10^{-1}$ | 2.33 | $7.75 \cdot 10^{-1}$ |
| | 31 | $2.43 \cdot 10^{-1}$ | $4.70 \cdot 10^{-1}$ | $3.84 \cdot 10^{-1}$ |

**Table 2.**
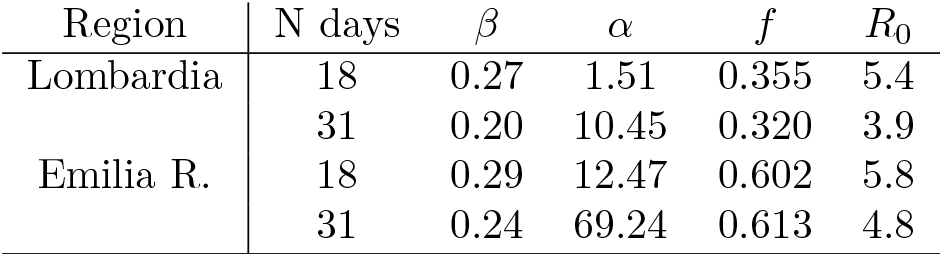
SEIRD Model parameters. Lombardia and Emilia Romagna regions

| Region | N days | $\beta$ | $\alpha$ | $f$ | $R_0$ |
| --- | --- | --- | --- | --- | --- |
| Lombardia | 18 | 0.27 | 1.51 | 0.355 | 5.4 |
|  | 31 | 0.20 | 10.45 | 0.320 | 3.9 |
| Emilia R. | 18 | 0.29 | 12.47 | 0.602 | 5.8 |
|  | 31 | 0.24 | 69.24 | 0.613 | 4.8 |

### 2.2 SEIRD(rm) calibration

In this paragraph we perform the calibration of the SEIRD(rm) model (1) where the coefficient *β* is assumed as the time dependent function (3). We perform a two phase process, where Phase 1 is carried out by running the classical SEIRD (1) for 18 days and Phase 2 applies the time dependent piecewise infection ratio *β*(*t*) in (3) all aver the measurements interval [*t*_0_, *t*_*p*_], partitioned as reported in table 3.

**Table 3.** Phase 2 parameters of *β*(*t*) in (3)

| Region | $t_0, t_1, \dots$ | $\rho_0, \rho_1, \dots$ |
| --- | --- | --- |
| Lombardia | [0, 10, 18, 24, 29] | [0, 0.9, 0.95, 0.999] |
| Emilia R. | [0, 7, 14, 21, 29] | [0, 0.2, 0.75, 0.9] |

In table 4 we report the relative errors obtained the SEIRD(rm) after calibration of SEIRD(rm) in the measurement interval [*t*_0_, *t*_*p*_]. Comparing the relative errors in the second and fourth rows of table 1 and 4, we can appreciate the improvement obtained by the calibrated model for lombardia region. Concerning Emilia Romagna only the Infected compartment reports a substantial improvement. We point out that while the behaviour of the Infected population is similar in the two regions, the Recovered and Dead are quite different, partly due to non homogeneous data acquisitions procedures in Lombardia and Emilia-Romagna.

**Table 4.** SEIRD(rm) Phase 2 Relative Errors

| Region | $Erel(I)$ | $Erel(R)$ | $Erel(D)$ |
| --- | --- | --- | --- |
| Lombardia | 5.60 $10^{-2}$ | 6.80 $10^{-2}$ | 7.75 $10^{-2}$ |
| Emilia R. | 5.40 $10^{-2}$ | 1.07 | 6.03 $10^{-1}$ |

The model parameters *α* and *f* obtained by this calibration phase are reported in table 5 and the functions *β*(*t*) are plotted in Fig. 5.

**Table 5.**
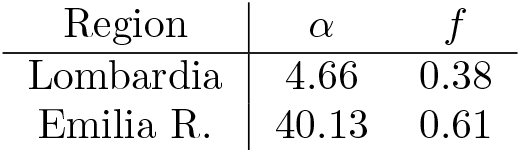
SEIRD(rm) parameters

| Region | $\alpha$ | $f$ |
| --- | --- | --- |
| Lombardia | 4.66 | 0.38 |
| Emilia R. | 40.13 | 0.61 |

**Fig 5.**
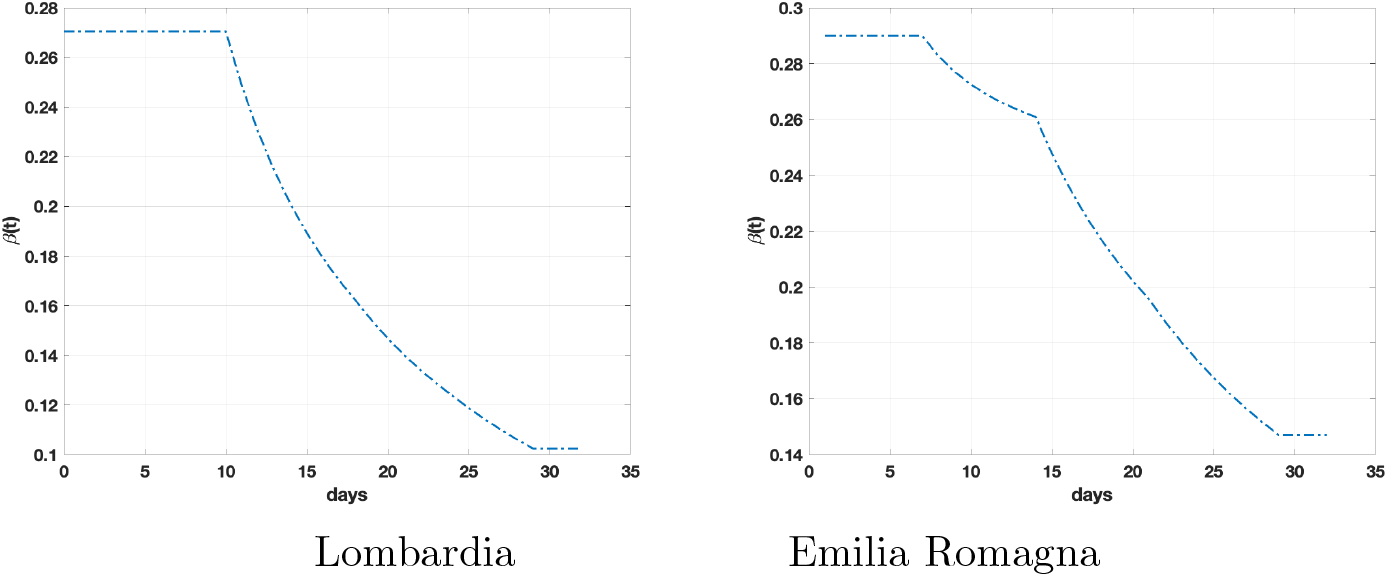
Infection Rate *β*(*t*) of SEIRD(rm) model.

We observe from the values of *R*_*t*_, reported in Fig.6, that *R*_*t*_ *>* 2 in both regions, meaning that the epidemic is still spreading, although less than in the first days.

**Fig 6.**
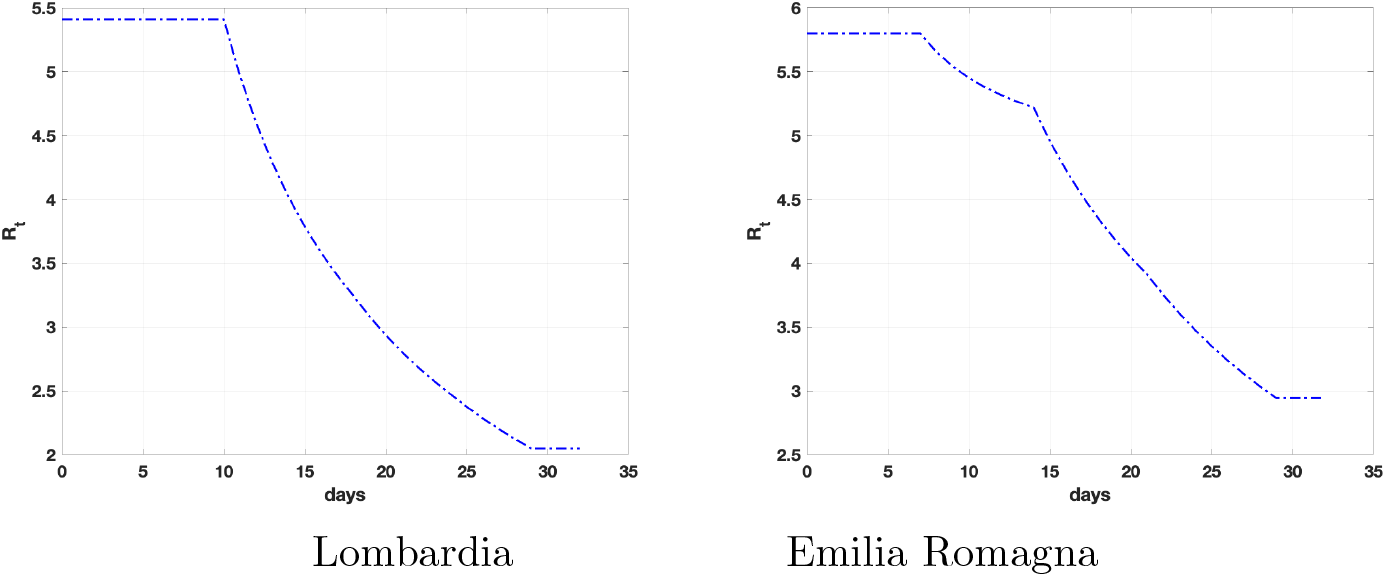
Reproduction Rate *R*_*t*_ of SEIRD(rm) model.

### 2.3 Epidemic Evolution forecasts

In this paragraph, we apply SEIRD(rm) to obtain information about the expected evolution of Infected, Recovered and Dead until 20/10/2020. We have chosen an Lombardia Emilia Romagna extended period for prevision, but we are aware that the effectiveness of the model decreases by getting away from the data.

In the following, we denote as [*t*_*p*_, *t*_*s*_] the prevision interval [28*/*3*/*2020, 20*/*10*/*2020]. Applying SEIRD(rm), calibrated in section 2.2, to the whole prevision interval, we obtain quite dramatic forecasts, as evident in table 6, where the Infected reach the maximum value around July 31 2020 (Lombardia) and June 4 2020 (Emilia Romagna), and there is an impressive number of deaths.

**Table 6.** SEIRD(rm) prediction of Infected, Recovered Dead maximum values over 240 days.

| region | Peak Infected |  | Peak Recovered |  | Peak Dead |  |
| --- | --- | --- | --- | --- | --- | --- |
|  | day | Number (%) | day | Number (%) | day | Number (%) |
| Lombardia | 149 | 1615056 (16.0%) | 240 | 4823468 (47.8%) | 240 | 2893530 (28.7%) |
| Emilia R. | 102 | 1305746 (29.3%) | 240 | 1750131 (39.2%) | 240 | 2411924 (54.1%) |

In Fig. 7, we can see, for example, the trend of Infected individuals in both regions. Although the actual SEIRD(rm) model fits the observations quite accurately, we believe that it does not reproduce the effects that the restrictions recently imposed, produce in the next future. Hence in the prediction interval [*t*_*p*_, *t*_*s*_] we change the model of the infection rate function, in the hypothesis that it continues to decrease.

**Fig 7.**
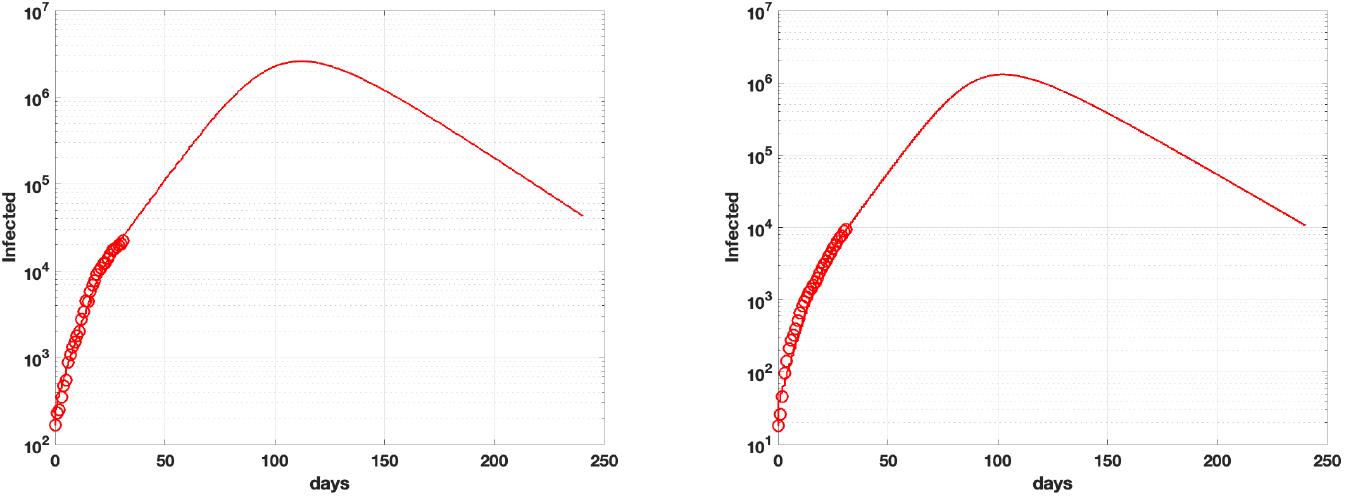
Infect trends SEIRD(rm) model.

In particular we show the results obtained by applying in the prevision interval [*t*_*p*_, *t*_*s*_] the SEIRD(rm) model with *α* and *f* as in table 5, and change *β*(*t*) with functions *β*_*r*_ (8), *β*_*c*_ (7) and *β*_*e*_ (9), with the value *β*_0_ set as *β*(*t*_*p*_), computed in the calibration phase 2.2. The *t*_*k*_ and *ρ*_*k*_ parameters for each formula are reported table 7.

**Table 7.** Prevision parameters of the infection rate in the interval [*t*_*p*_, *t*_*s*_.

| function | $t_0, t_1, \dots$ | $\rho_0, \rho_1, \dots$ |
| --- | --- | --- |
| $\beta_r$ | [32, 59, 240] | [0.9999, 0.999999] |
| $\beta_c$ | [32, 46, 60, 240] | [0.75, 0.55, 0.30, 0.20] |
| $\beta_e$ | [32, 46, 60, 240] | [0.05, 0.07, 0.09, 0.1] |

#### 2.3.1 Emilia Romagna Forecasts

From results in table 8, we observe that the piecewise exponential model (function *β*_*e*_) gives the smallest number of Infected-Recovered-Dead individuals.

**Table 8.** SEIRD(rm) Emilia Romagna prediction, Infected, Recovered Dead maximum values over 240 days

| Data<br>$\beta(t)$ | Peak Infected | | Peak Recovered | | Peak Dead | |
| --- | --- | --- | --- | --- | --- | --- |
|  | day | Number (%) | day | Number (%) | day | Number (%) |
| $\beta_r$ | 92 | 76815 (1.7%) | 240 | 169493 (3.8%) | 240 | 233586 (5.2%) |
| $\beta_c$ | 60 | 37143 (0.8%) | 240 | 58418 (1.3 %) | 240 | 80509 (1.8%) |
| $\beta_e$ | 52 | 25581 (0.6%) | 240 | 26298 (0.6%) | 240 | 36242 (0.8%) |

These results represent the best situation, where the restriction measures have the strongest impact. The epidemic peak is expected on April 15th (Fig. 8). The piecewise constant decreasing *β*_*c*_ has a slightly different result, with the maximum Infections reached on the first week of April 2020 (Fig. 10). The piecewise rational model *β*_*r*_ provides the most pessimistic prevision, with about 77000 infectious people and more than 233000 dead (Fig. 9). Concerning the epidemic spread we can appreciate the different behaviours of the basic reproduction function *R*_*t*_ in Fig. 11. The

**Fig 8.**
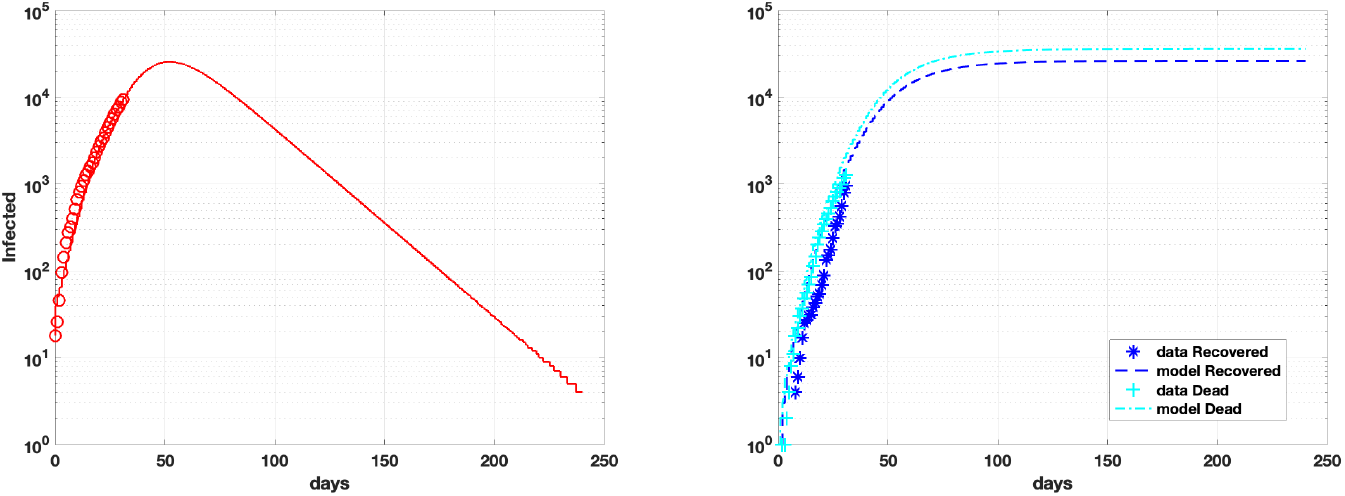
Emilia R. forecast Infect-Recovered-Dead, piecewise exponential model.

**Fig 9.**
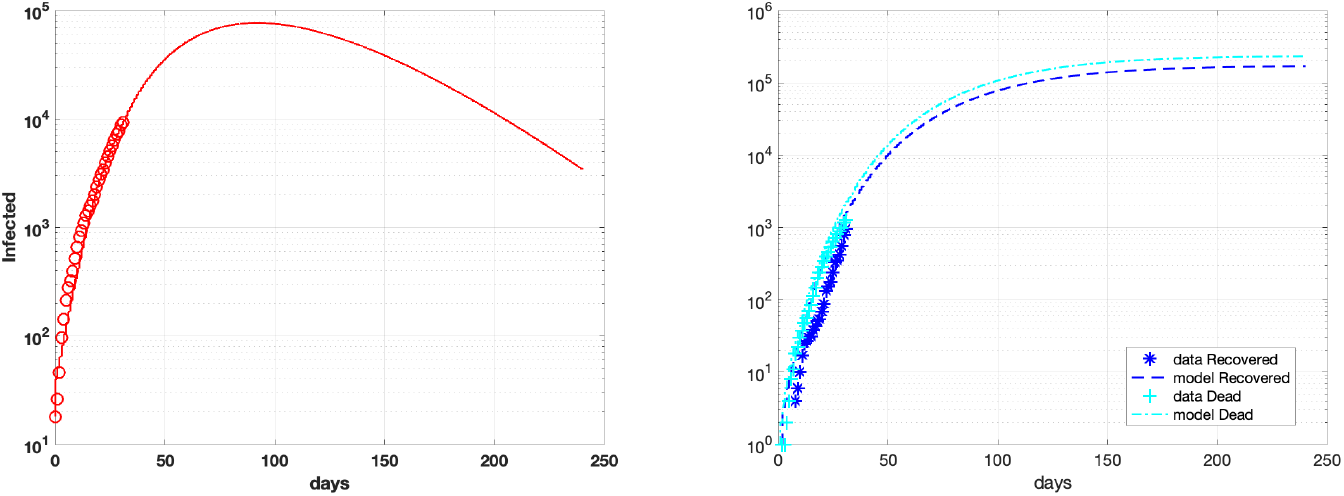
Emilia Romagna forecast Infect-Recovered-Dead, piecewise rational model.

**Fig 10.**
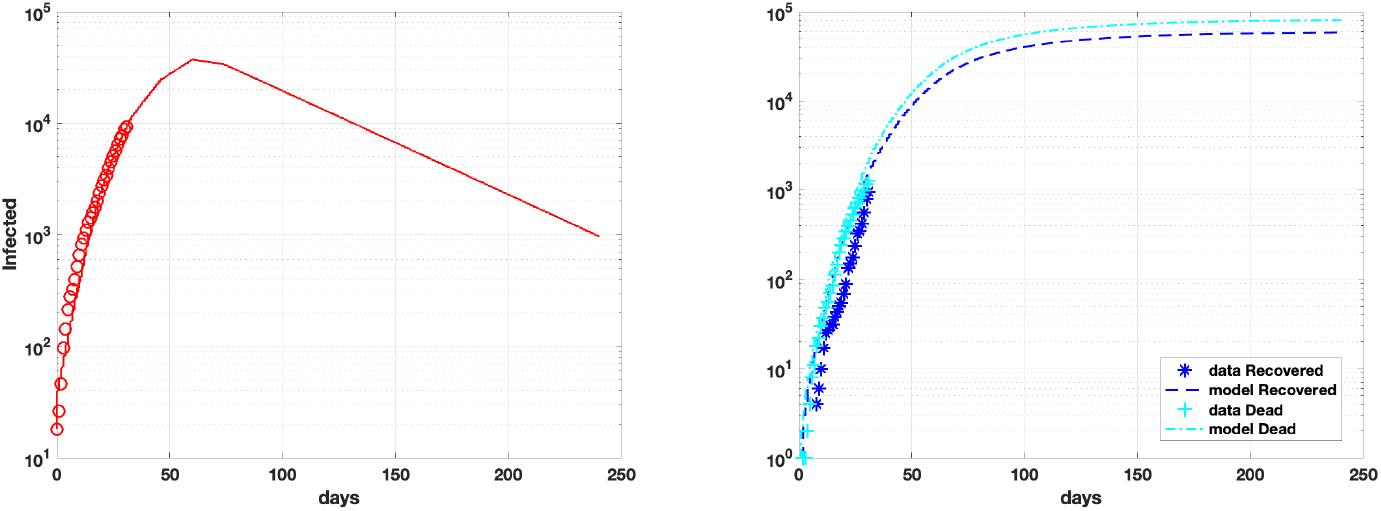
Emilia R. forecast Infect-Recovered-Dead, piecewise constant model.

**Fig 11.**
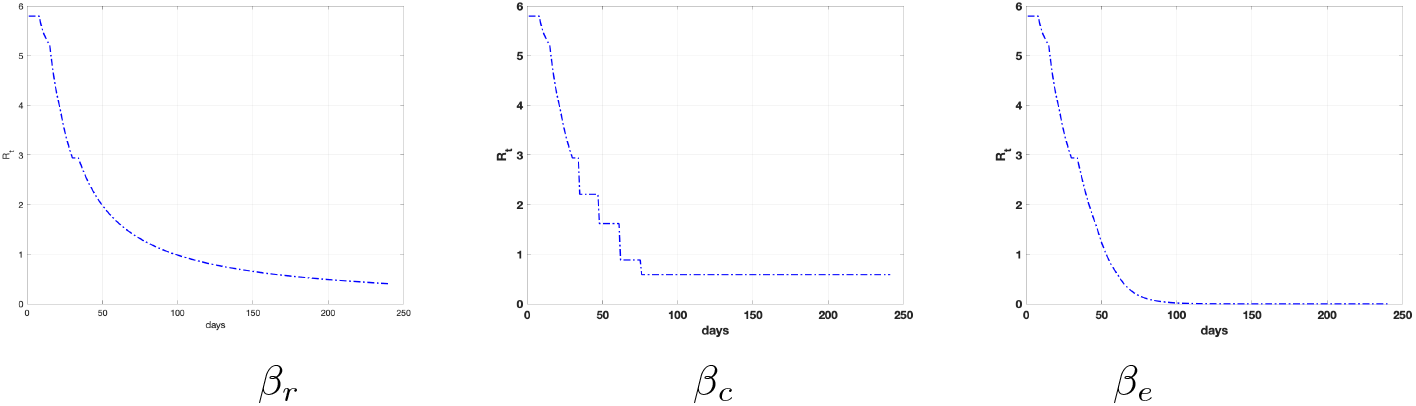
Emilia Romagna *R_t* functions obtained with SEIRD(rm) and *β*_*r*_ (8), *β*_*c*_ (7) and *β*_*e*_ (9) in the prediction interval [*t*_*p*_, *t*_*s*_].

#### 2.3.2 Lombardia forecasts

In table 9 we report the peak values of the Infected-Recovered-Dead populations obtained with the proposed SEIRD(rm) model relative to the Lombardia region. Table 9 shows that again the piecewise exponential model predicts the peaks sooner than the others and with the lowest number of Infected, Recovered and Dead (see figures Fig. 15, 14, 13). The behaviour of the epidemic spread is reported in Fig. 12.

**Table 9.** SEIRD(rm) Lombardia prediction, Infected, Recovered Dead maximum values over 240 days

| $\beta(t)$ | Peak Infected | | Peak Recovered | | Peak Dead | |
| --- | --- | --- | --- | --- | --- | --- |
| $\beta$ | day | Number (%) | day | Number (%) | day | Number (%) |
| $\beta_r$ | 67 | 47257 (0.5%) | 240 | 139389 (1.4%) | 240 | 83617 (0.8%) |
| $\beta_c$ | 60 | 36668 (0.4%) | 240 | 93388 (0.9%) | 240 | 56022 (0.6%) |
| $\beta_e$ | 47 | 33277 (0.3%) | 240 | 54135 (0.5%) | 240 | 32475 (0.3%) |

**Fig 12.**
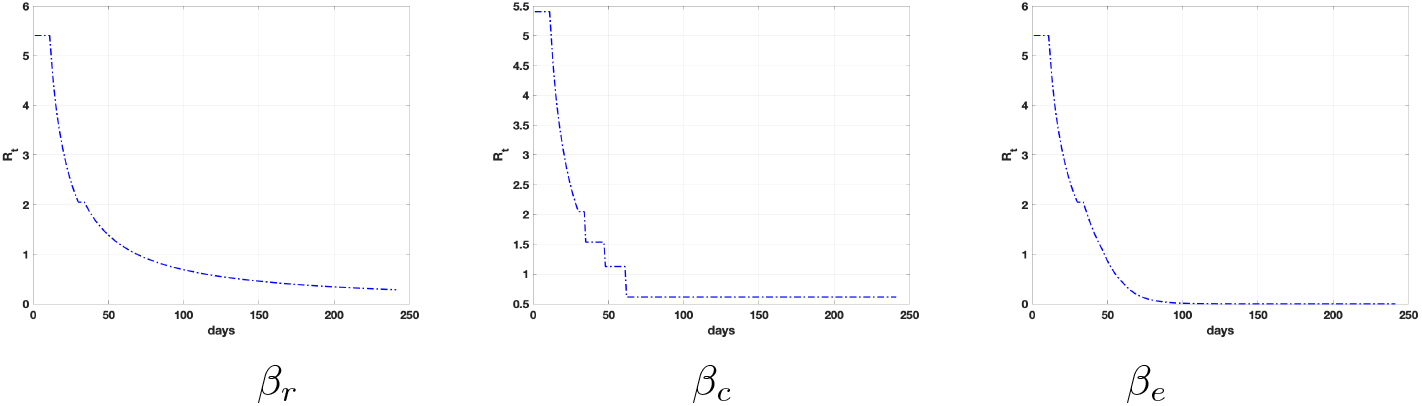
Lombardia *R_t* functions obtained with SEIRD(rm) and *β*_*r*_ (8), *β*_*c*_ (7) and *β*_*e*_ (9) in the prediction interval [*t*_*p*_, *t*_*s*_].

**Fig 13.**
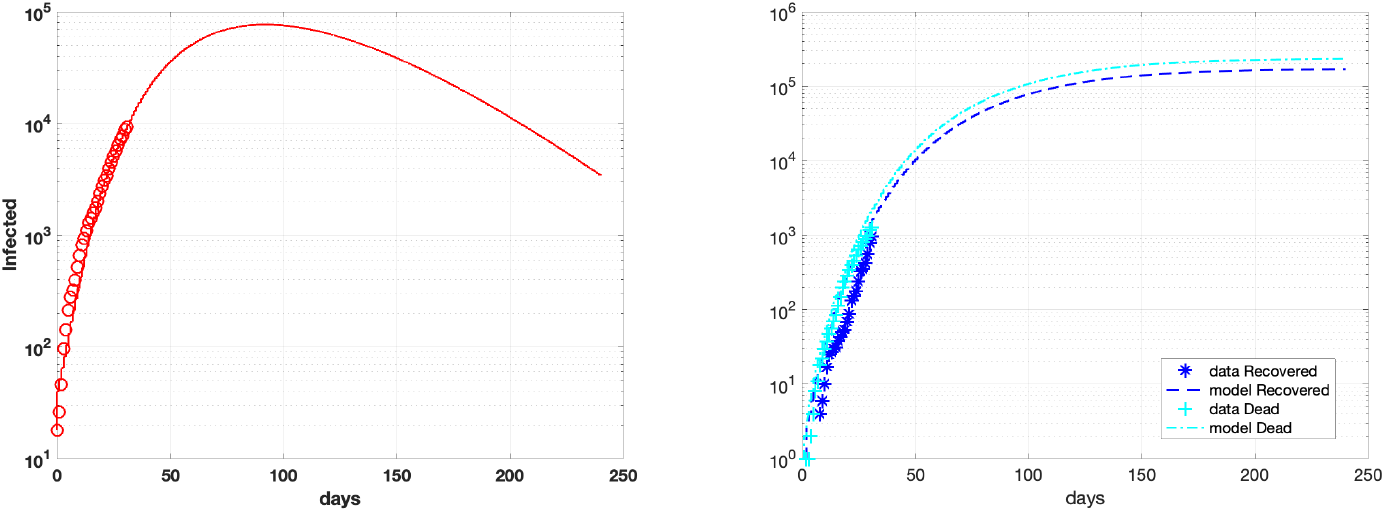
Lombardia forecast Infect-Recovered-Dead, piecewise rational model *β*_*r*_.

**Fig 14.**
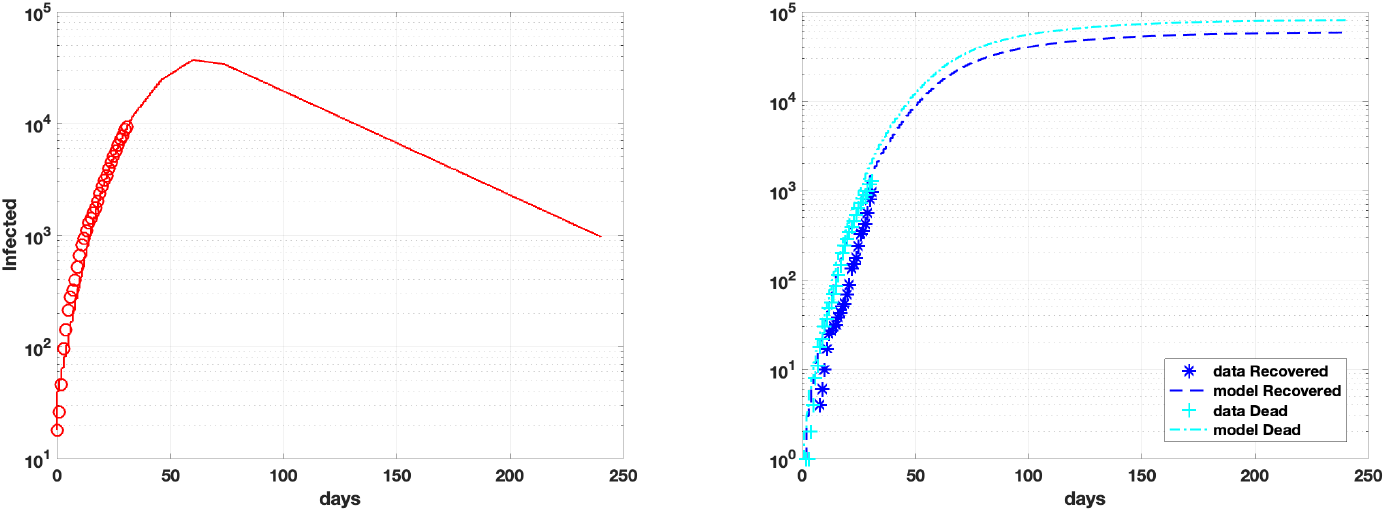
Lombardia forecast Infect-Recovered-Dead, piecewise constant model*β*_*c*_.

**Fig 15.**
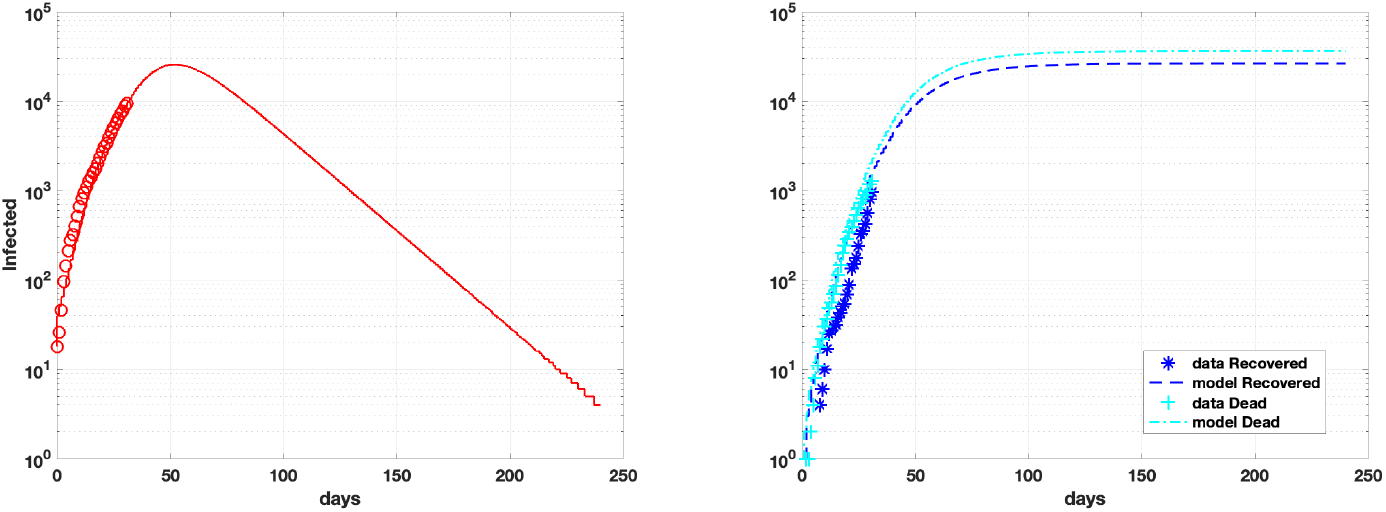
Lombardia forecast Infect-Recovered-Dead, piecewise exponential model *β*_*e*_.

## Conclusion

In this paper, we proposed a SEIRD model for the analysis of the COVID-19 outbreak diffusion in Italy. In our new formulation, we have partitioned the integration time into subintervals, where the infection rate coefficient has been adaptively modelled as a decreasing function of the time. This model flexibly takes account of the successive lockdown restrictions imposed by the Italian Government to contain the outbreak. The results obtained by fitting the data of two Italian regions, Lombardia and Emilia-Romagna, available since February 24th 2020, show a very good fit to the data, with very small errors.

We have then used the model to forecast the epidemic spread for future times. We have considered three different time-dependent functions to describe the infection rate coefficient and shown the simulation results obtained with each of them in Lombardia and Emilia-Romagna.

We highlight that only a few days passed since restrictions started in Italy and, maybe in the next few days, the effects of such measures will be more evident, hopefully causing a further decrease in the infection trend. In this case, the previsions shown in this paper should be updated by introducing a new time intervals at which the decreasing slope of *β*_*t*_ should be easily changed by simply modifying one parameter.

The proposed model is flexible and can be quickly adapted to monitor various infected areas with different restriction policies. Moreover, the parameters *ρ*_*i*_, now fixed, as in table (3), could be estimated in the future by an identification procedure.

## Data Availability

The code is available upon request to the authors.

## References

1. Mathematical epidemiology: Past, present, and future. Infectious Disease Modelling. 2017;2(2):113–127. doi: https://doi.org/10.1016/j.idm.2017.02.001.

2. A primer on stochastic epidemic models: Formulation, numerical simulation, and analysis. Infectious Disease Modelling. 2017;2(2):128–142. doi:https://doi.org/10.1016/j.idm.2017.03.001.

3. O Kw, G Ma. A contribution to the mathematical theory of epidemics. In: Proceedings of the Royal Society of London. vol. A; 1927. p. 700–721.

4. Boujakjian H. Modeling the Spread of Ebola with SEIR and Optimal Control. In: SIAM Under-graduate Research Online. 9. SIAM; 2016. p. 299–310.

5. Chowell G, Hengartner N, Castillo-Chavez C, Fenimore P, Hyman J. The basic reproductive number of Ebola and the effects of public health measures: the cases of Congo and Uganda. Journal of Theoretical Biology. 2004;(229):119–126.

6. Kucharski A, Russell TW, Diamond C, Liu Y, Edmunds J, Funk S, et al. Early dynamics of transmission and control of COVID-19: a mathematical modelling study. Lancet. 2020;doi: https://doi.org/10.1016/S1473-3099(20)30144-4.

7. Wu J, Leung K, Leung G. Nowcasting and forecasting the potential domestic and international spread of the 2019-nCoV outbreak originating in Wuhan, China: a modelling study. Lancet. 2020;(395):689–697. doi: https://doi.org/10.1016/S0140-6736(20)30260-9.

8. Tanga Z, Lib X, Lic H. Prediction of New Coronavirus Infection Based on a Modified SEIR Model;.

9. Repository D. https://github.com/pcm-dpc/COVID-19;.

10. S WJ, J D. Modeling post-death transmission of Ebola: challenges for inference and opportunities for control. Scientific reports. 2015;(5: 8751). doi:10.1038/srep08751.

11. Zama F, Frascari D, Pinelli D, Bacca AEM. Parameter Estimation Algorithms for Kinetic Modeling from Noisy Data. In: System Modeling and Optimization. Springer International Publishing; 2016. p. 517–527.

